# A Roadmap to Artificial Intelligence (AI): Methods for Designing and Building AI ready Data for Women’s Health Studies

**DOI:** 10.1101/2023.05.25.23290399

**Authors:** Farah Kidwai-Khan, Rixin Wang, Melissa Skanderson, Cynthia A. Brandt, Samah Fodeh, Julie A. Womack

## Abstract

**Objectives:** Evaluating methods for building data frameworks for application of AI in large scale datasets for women’s health studies.

**Methods:** We created methods for transforming raw data to a data framework for applying machine learning (ML) and natural language processing (NLP) techniques for predicting falls and fractures.

**Results:** Prediction of falls was higher in women compared to men. Information extracted from radiology reports was converted to a matrix for applying machine learning. For fractures, by applying specialized algorithms, we extracted snippets from dual x-ray absorptiometry (DXA) scans for meaningful terms usable for predicting fracture risk.

**Discussion:** Life cycle of data from raw to analytic form includes data governance, cleaning, management, and analysis. For applying AI, data must be prepared optimally to reduce algorithmic bias.

**Conclusion:** Algorithmic bias is harmful for research using AI methods. Building AI ready data frameworks that improve efficiency can be especially valuable for women’s health.

**Lay Summary:** Women’s health studies are rare in large cohorts of women. The department of Veterans affairs (VA) has data for a large number of women in care. Prediction of falls and fractures are important areas of study related to women’s health. Artificial Intelligence (AI) methods have been developed at the VA for predicting falls and fractures. In this paper we discuss data preparation for applying these AI methods. We discuss how data preparation can affect bias and reproducibility in AI outcomes.

## Background

### Clinical Significance

Falls and fractures are important outcomes to study for women’s health. Here we evaluate two studies where falls and fractures are studied amongst United States Veterans who present to the VA for care. There are over 13 million Veterans in care with more than 900,000 women Veterans. Studies have shown model bias in AI can impact diagnosis in clinical care.^20^ Medications such as bisphosphonates are used to reduce fracture risk by as much as 25% to 50%.^4,10,11^ However they can also have side effects,^6,7^ especially in people with comorbid conditions. Accurately identifying patients who would benefit from bisphosphonate therapy is an important and complex clinical decision. In 2008, investigators developed and validated the Fracture Risk Assessment tool (FRAX), an algorithm which calculates the 10-year probability of hip and major-osteoporotic fracture (clinical vertebral, hip, forearm, or upper arm). FRAX includes modifiable and non-modifiable risk factors: age, sex, alcohol and tobacco use, prior osteoporotic fracture, history of parental fracture, body mass index (BMI), rheumatoid arthritis, oral glucocorticoid use, and secondary causes of osteoporosis, with or without femoral neck bone mineral density (FN)-BMD.^5^ However, it underestimates fracture risk among people with chronic conditions such as HIV^8,9^.

Previous studies have shown women are at a higher risk of bone density loss with age^18^ than men. With an evolution in tools for predicting disease, we need additional research to apply these methods for women’s health. The goal of this study was to prepare data with a reproducible and rigorous approach for foundational work required to create machine learning algorithms that reliably predict risk of falls and use NLP for calculation of T-score for predictive fractures amongst Veterans.

### Informatics Significance

Understanding the context of data collection is foundational to research^19^. We may risk misinterpreting data if we do not factor in its provenance. We may even introduce bias if the underlying data misrepresents sample characteristics such as race, prevalence of disease or clinical outcomes.^20^ Literature shows the most important application of machine learning to health data so far has been phenotyping, classifying and categorizing patients with a disease, as patients can be retrospectively selected using already existing data. Foundational work is required to prepare electronic health record data along with unstructured text notes in a reproducible manner that is based on data structuring principles with continuous vigilance of algorithms for fairness^20^.

Data structure may be based on the practices and policies of the organization. In structuring data, the organizational practices may need to be addressed with policy makers. Insights into why data might be sparse for a specific group or how to handle missing data must be carefully integrated within data cleaning and structuring techniques. Data frameworks that address bias and improve reproducibility are essential for building fair, explainable AI models^20^. The aim of this study is to evaluate application of carefully curated methods for preparing electronic health record data to be the foundation for machine learning algorithms for predicting risk of fracture and identifying falls. There have been studies that explore the ability of machine learning to make predictions on falls^13^, here we focus on the data preparation methodology that the machine learning algorithms are based on.

## Methods

### Data Structuring

We evaluated data preparation methods utilized for two studies assessing risk of falls and fractures amongst Veterans. For the falls study we created a cohort of people who had a serious fall (significant enough to result in a visit to a healthcare provider) identified by international classification codes and those who did not have a diagnosis code. We then created a database with data from different domains of the electronic health record. The domains included pharmacy, visits, diagnosis, demographic, radiology, and progress notes. We applied big data management and programming techniques to build the cohort. The VA data includes millions of patients with records across multiple domains that need extract load transform (ETL) routines to create cohorts of interest. On these data, programming done in structured query language (SQL) and statistical analysis system (SAS) was applied to create clinically meaningful variables. These variable definitions have been validated over a period across multiple studies.^14,15^ Figure1 represents a simplified depiction of the flow of data from the corporate data warehouse (CDW) through SQL table joins for the relevant domains using application of programmatic routines. The performance of these routines was optimized based on the processing power of the environment. The VA’s informatics and computing infrastructure (VINCI) is a unique environment available to researchers with real time electronic health record data with high efficiency computing power.

**Figure 1:**
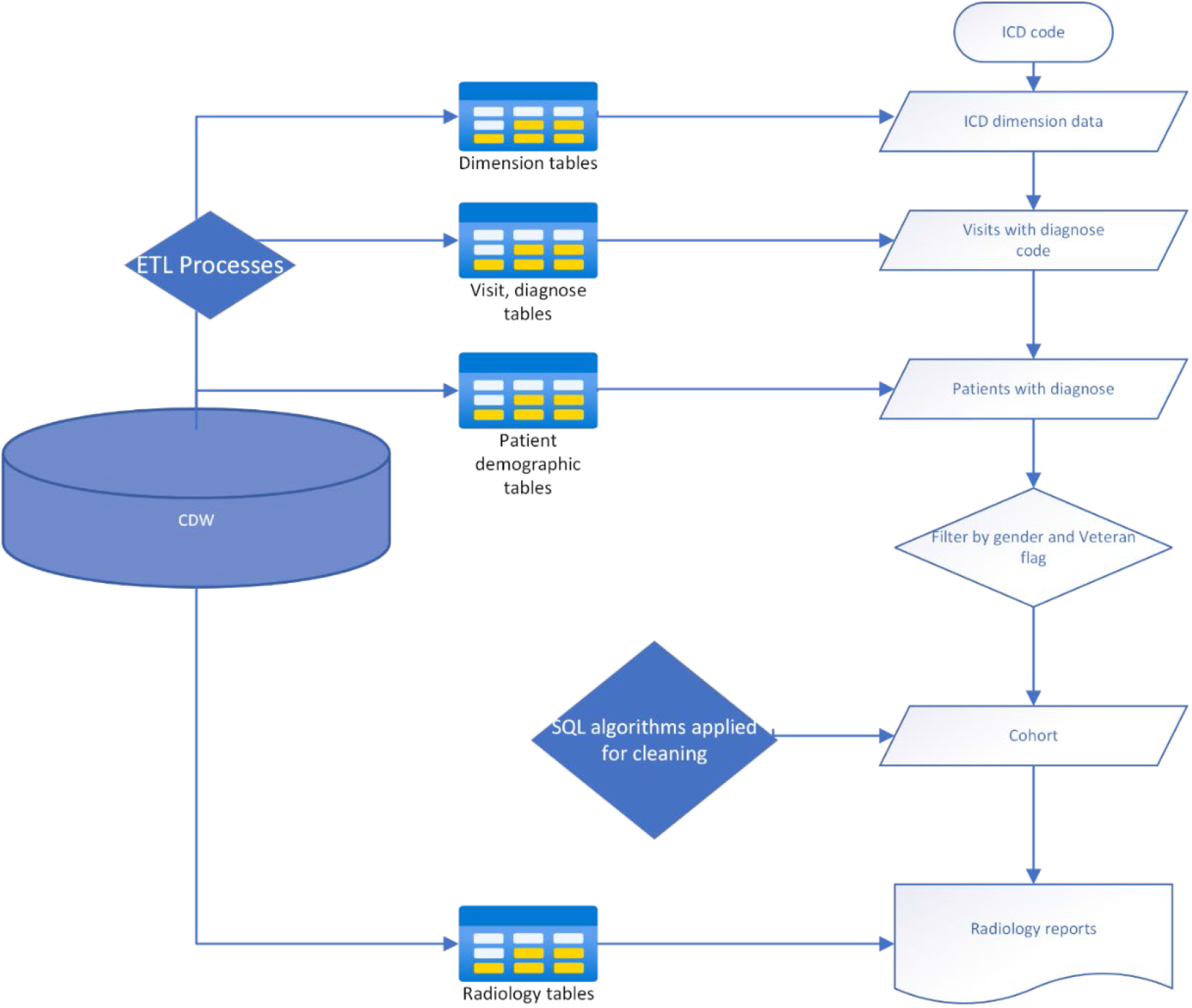
Data collection process

### Implementing a rule based algorithm

Structured electronic health record data often lacks concepts that may be identifiable using AI methods. Clinical notes are free text or unstructured data. On the notes we followed specialized data preparation techniques with natural language processing (NLP) for identifying falls. There are no standard entries, no numerical result to be filled into a field. Clinicians may use different sentences and words to describe the same concept. Variations include word order, word variants, abbreviations, acronyms, synonyms, punctuation, misspelling etc. Therefore, NLP was an important step before we could feed clinical notes into machine learning algorithms. Data representation was the first and most critical task towards building the classifier. For the study analysis a machine learning model was developed to identify the fall event for radiology report, in which each report was represented by a feature set of words and Unified Medical Language System (UMLS) concepts.

We utilized YTEX, an NLP tool built on top of Apache clinical Text Analysis and Knowledge Extraction System (cTAKES), which comprises sentence splitting, tokenization, part-of-speech tagging, shallow parsing, named entity recognition, and storage of all annotations in a database. The CTAKES/YTEX application created normalized output of the tokenizer and the concept unique identifiers of the named entity recognition components and saved in database tables. The tokenizer component included word tokenization, normalization (lowercase, stemming, and lemmatization), stop words removal. The named entity recognition component maps span of text to a dictionary lookup table, which has been prepopulated with UMLS concepts based on the study subject. For this study, a combination of words and concept identifiers was used as the feature. Each radiology report as a “bag of features” was represented by a binary feature vector. The extracted information from radiology reports was converted into a structured form (matrix) which the machine learning techniques were applied. Figure 2 illustrates the flow mentioned above in a graphic format.

**Figure 2:**
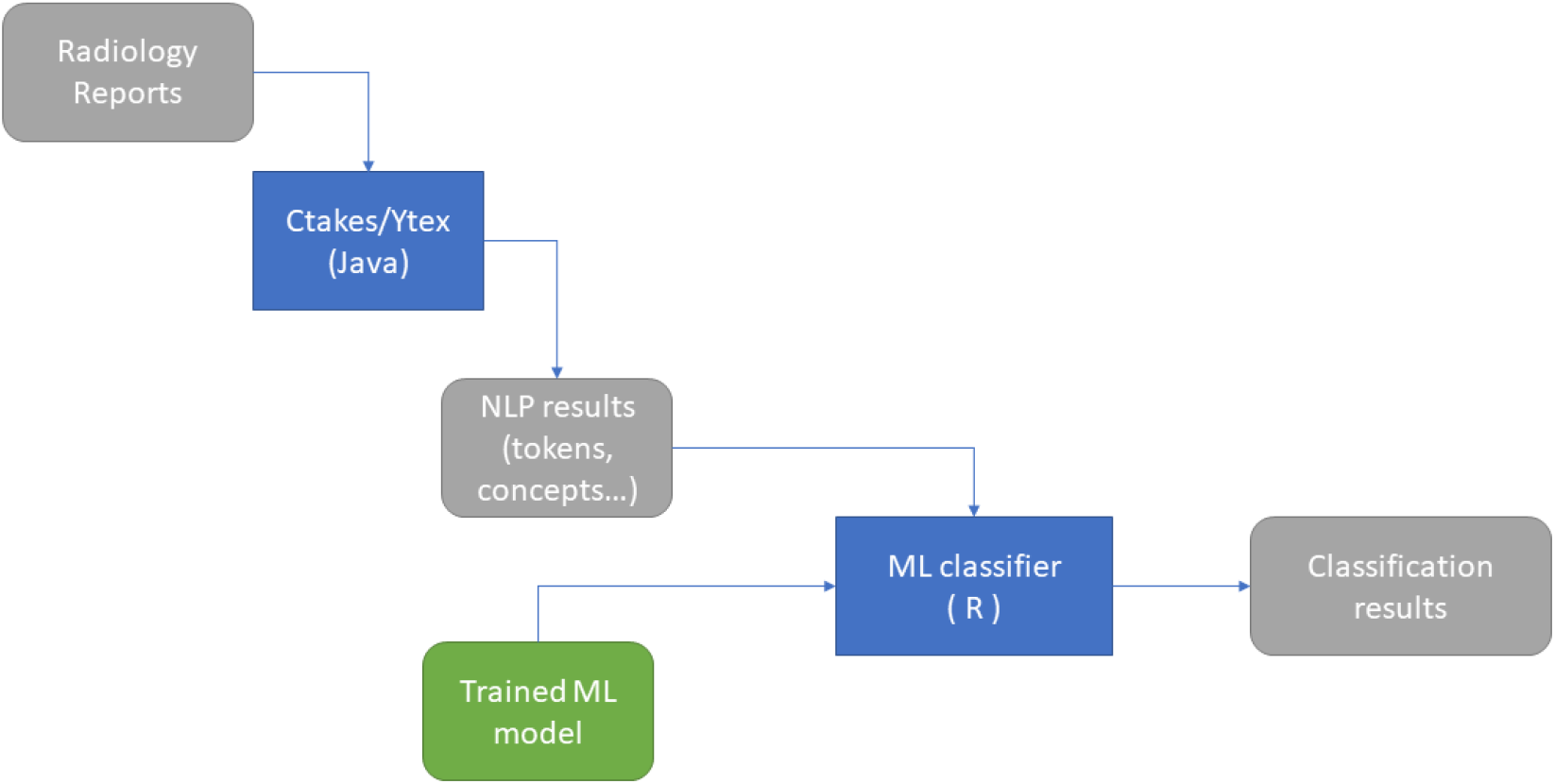
Falls classification workflow

### Snippet formulation

For the fractures study, the cohort database includes different domains of the electronic health record such as pharmacy, visits, diagnosis and notes from both outpatient and inpatient categories. Data was stratified by age, race, and gender. Natural language processing methods were run on the DXA scan reports to create snippets for creating context for a predictive model using machine learning techniques. Big data management and querying techniques were applied by utilizing SQL Server analysis, .NET and other data warehousing and management tools. For getting the data ready for running the machine learning algorithms, we used international procedure codes (CPT) from procedures to identify DXA scans. Once people with the relevant procedure codes were identified, we collected 1,387,479 radiology reports related to these scans from unstructured radiology exam tables. From these report texts we created snippets of text related to key terms identified by clinicians that would be relevant in mining these reports for additional context. These snippets were generated with specialized algorithms to extract relevant context for the risk prediction analysis. Multiple iterations are typically required of the snippet extraction process to provide enough context in the text for the machine learning algorithms. We did chart reviewed on a set of notes to rule out possibility of irrelevant imaging data. SQL functions were utilized to perform extraction of the snippets for relevant keywords. The coding to extract snippets from unstructured text was done by creating specialized SQL functions. The functions were called within queries that were applied in pulling notes with the relevant snippets. The routines were created with generalizability to run multiple iterations for different snippet sizes. This helped to optimize snippet size for the NLP algorithms. Figure 3 represents a simplified view of the basic steps involved in getting data ready for the machine learning algorithm. The base of the pyramid is the data collection followed by creation of relevant objects.

**Figure 3:**
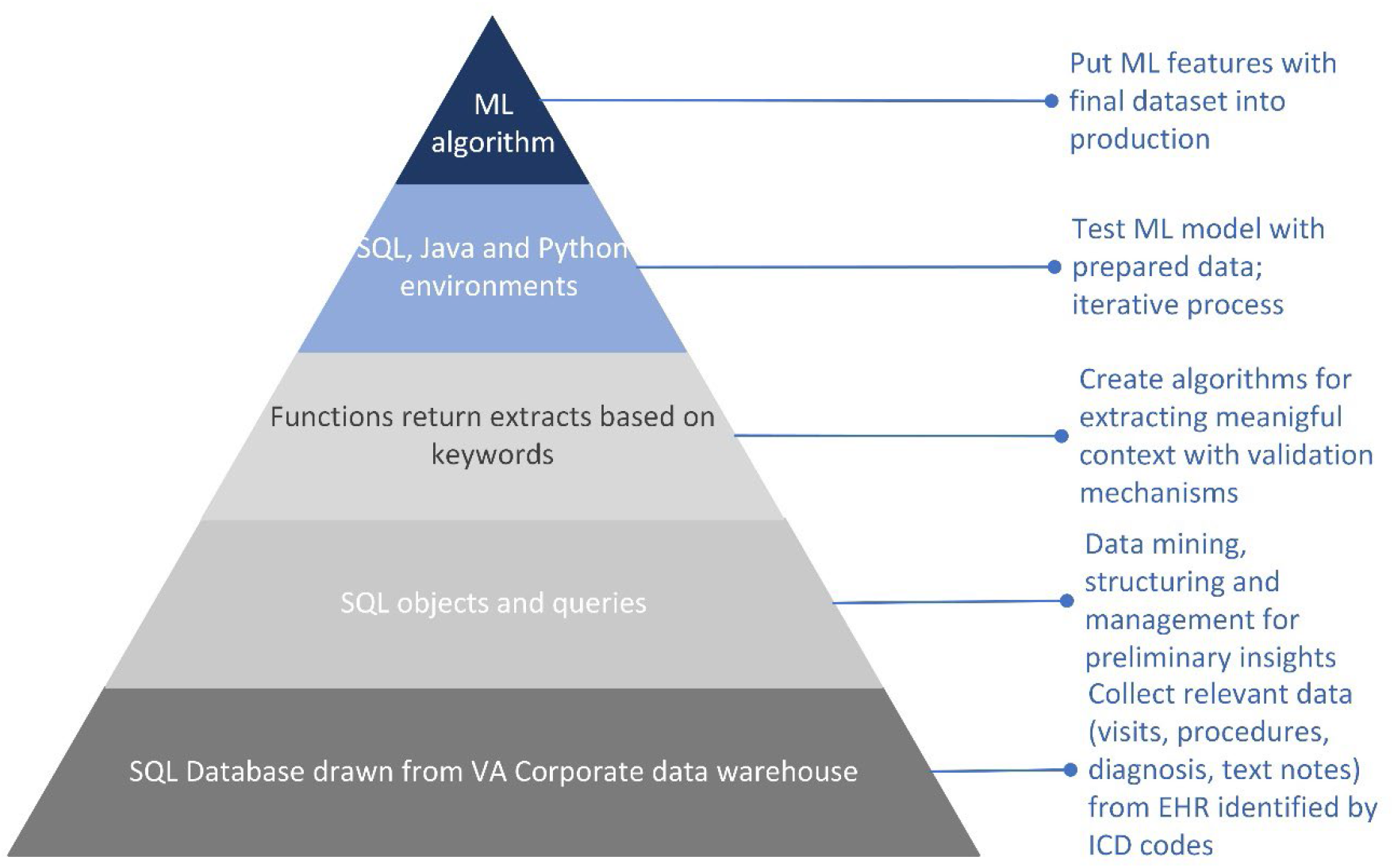
Data preparation pyramid

For the fractures study, T-score (bone density comparison to normal value) was a key factor in calculating risk analysis. To prepare data for NLP, we developed a rule-based algorithm to retrieve the T-Score from the snippets. In the radiology report with DXA scan, the T-score presents around the keyword with two possible formats, in data table or in plain language.

We developed two group of regular expressions, one group is used to retrieve the T-score values in table format, in this case, we first match the table header in snippet, then match the value in following context. Another group of regular expression is used to match the T-score in plain language. In radiology notes, the T-score always presented with sentence patterns. We annotated many snippets with T-score in plain language before developing the pattern match regular expressions.

## Results

For Falls, 5,377,673 reports were processed by the machine learning algorithm out of which 45,304 were flagged as a positive prediction and 5,332,369 as negative. Table 1 summarizes the output by gender. Amongst a cohort of 13 million Veterans (917,596 women), 695,645 Veterans (159,454 women) had DXA imaging from 2000 to 2020. There were 1,387,479 radiology reports associated with these visits. For the DXA analysis we found that the length of the snippet is an important factor for match accuracy. Shorter snippet length will miss important sentence parts and lead to missed T-score values. Longer snippet length was desirable, but if snippets are too long, they generate some false T-score matches. After we tested different length of the snippets (including whole report), we found the snippets with 50 words before and after keyword is a proper length. The final predictive model for fragility fractures study is being finalized using the foundational data framework discussed earlier. So far, NLP algorithms have yielded 95% correct T-score values from snippets (recall 98%, error rate 1.9%).

**Table 1:** Prediction output

| Radiology Reports | Prediction | Percent prediction (%) |
| --- | --- | --- |
| **Female** |  |  |
| 1528 | Negative | 84 |
| 285 | Positive | 16 |
| **Male** |  |  |
| 51934 | Negative | 89 |
| 6277 | Positive | 11 |

## Discussion

Algorithmic bias can introduce inequity in clinical decision making and healthcare research^1,12,16^. To prevent bias and produce reliable results, the data that are the foundation for the algorithms need careful curation especially in women’s health^17,20^. This implies data readiness for AI is not a one size fits all solution^2,3^. The preparation of data depends on the specific AI method and the context of the research question’s data domain.

With healthcare data, the interaction of elements such as data governance, cleaning, management, analysis, and organizational policies should be carefully assessed when applying AI methods. Structuring input to AI algorithms in ways relevant to the specific AI method being utilized creates a wholistic approach to address the research question. The data preparation methods we evaluated and applied in this study directly synergized with the AI algorithms. For future work we propose keeping provenance of data on the forefront when deciding on methodologies for data curation.

## Conclusion

The roadmap of data for application of AI involves applying specialized techniques to develop a framework that promotes transparency and improves efficiency. Algorithmic bias can be harmful for women’s health studies, if not carefully evaluated. Therefore, building AI ready data frameworks and establishing the informatics processes to do so can help promote equity in decision making and healthcare research.

## Data Availability

Data from this study is behind the VA firewalls and can be obtained with necessary approvals upon request

## Acknowledgements

The authors acknowledge the Veterans without whom the data at the VA would not be available for this research. The views and opinions expressed in this manuscript are those of the authors and do not necessarily represent those of the Department of Veterans Affairs or the United States government. We have no conflicts or competing interests in the preparation of this manuscript.

